# Consensus study of risk factors and symptoms of SARS-CoV-2 (COVID-19) using biomedical literature and social media data

**DOI:** 10.1101/2020.05.17.20104729

**Authors:** Jouhyun Jeon, Gaurav Baruah, Sarah Sarabadani, Adam Palanica

## Abstract

**Background:** In December 2019, Coronavirus disease 2019 (COVID-19) outbreak started in China and rapidly spread around the world. Lack of any vaccine or optimized intervention raised the importance of characterizing risk factors and symptoms for the early identification and successful treatment of COVID-19 patients.

**Methods:** We systematically integrated and analyzed published biomedical literature and public social media data to expand our landscape of clinical and demographic variables of COVID-19. Through semantic analysis, 45 retrospective cohort studies, which evaluated 303 clinical and demographic variables across 13 different outcomes of COVID-19, and 84,140 tweet posts from 1,036 COVID-19 positive users were collected. In total, 59 symptoms were identified across both datasets.

**Findings:** Approximately 90% of clinical and demographic variables showed inconsistency across outcomes of COVID-19. From the consensus analysis, we identified clinical and demographic variables that were specific for individual outcomes of COVID-19. Also, 25 novel symptoms that have been not previously well characterized, but were mentioned in social media. Furthermore, we observed that there were certain combinations of symptoms that were frequently mentioned together among COVID-19 patients.

**Interpretation:** Identified outcome-specific clinical and demographic variables, symptoms, and combinations of symptoms may serve as surrogate indicators to identify COVID-19 patients and predict their clinical outcomes providing appropriate treatments.

**Funding:** This research was internally funded and received no specific grant from any funding agency in the public, commercial, or not-for-profit sectors.

## Introduction

Coronavirus disease 2019 (COVID-19) is an emerging infectious disease that has quickly spread worldwide. Since its outbreak in China in December 2019, over 4 million cases have been confirmed across more than 200 countries^1^ (as of May 15, 2020), with the number of cases continuing to increase. Several studies have characterized possible symptoms and risk factors for clinical outcomes^2,3^. However, the majority of retrospective studies have been based on a single center and counted the number of aggregate cases^4^, providing a scattered and incomplete picture of the risk factors for disease severity. Furthermore, uncharacterized or uncommon symptoms have made COVID-19 difficult to diagnose, and make it difficult to provide appropriate treatment to patients. Lack of vaccine and optimized treatment raise the importance of early and definitive diagnosis for this disease. Additionally, there are limited hospital resources to triage patients based on symptoms to determine who is more or less likely to require intensive treatment (*e.g*. ICU admission or intubation).

All of these uncertainties suggest there are urgent needs for a low-cost and efficient method of gathering COVID-19 symptom- and risk factor-related data as quickly as possible to reduce the medical and economic burden in our society. Instead of conducting time-consuming and costly clinical trials to examine patients, an alternative research avenue involves scraping public social media data to investigate potential clinical and demographic variables of COVID-19 development. Social media provides an efficient method of gathering large amounts of representative data on the general public, in a cost-effective, scalable, and convenient manner any time of day, especially in remote or unattended regions. Here, we systematically integrated and analyzed published biomedical literature and social media data to identify novel clinical and demographic variables specific for different outcomes of COVID-19, and also examined rare or uncommon symptoms that were associated with COVID-19 progression but were not previously well characterized.

## Methods

### Compiling biomedical literature of COVID-19, and identifying clinical and demographic variables

CORD-19 (COVID-19 Open Research Dataset)^5^ was used to find biomedical literature for COVID-19. We compiled retrospective studies that investigated clinical and demographic variables in various outcomes of COVID-19. To do this, we collected literature published between January, 2020 and March, 2020. We then generated two sets of keywords. The first set of keywords represented cohort-based and retrospective studies. Keywords were “epidemiological characteristics”, “clinical characteristics”, “risk factors”, “clinical features”, “cohort”, “clinical course”, “clinical findings”, “risk of death”, “pathological characteristics”, “retrospective”, and “mortality risk”). The second set represented COVID-19 (“novel coronavirus”, “coronavirus”, “COVID-19”, “SARS-COV-2”, “severe acute respiratory syndrome coronavirus 2”, “2019 novel coronavirus”, “2019-ncov”, and “coronavirus disease 2019”). From the semantic analysis, we found 116 articles that contained both keywords, and were likely to be relevant to our study. We next extracted 535 tables in those articles using Camelot^6^ based on the notion that tables listed clinical and demographical variables, and their associated statistics. After careful manual curation of tables, reported data, and article themselves, 45 studies were chosen (**Appendix 1**). From the literature, 304 clinical and demographic variables were characterized. They were composed of 92 comorbidities/complications, 49 treatments, 124 lab findings, 34 symptoms, and 4 demographic variables (age, sex, alcohol drinking history, and smoking history).

### Compiling social media data and identifying symptoms of COVID-19 positive users from social media data

Twitter was used as the social media source. To identify COVID-19 positive users, we first collected users who used one of the phrases: “my positive COVID test”, “my positive COVID diagnosis”, “I am positive for COVID”, “I tested positive for COVID”, and “I have COVID-19” in their tweets between January, 2020 and March, 2020. In total, 1,036 users were identified as COVID-19 positive users. We then collected an additional tweet posts generated 14 days before and 14 days after their original tweets mentioning COVID-19 status (84,140 tweets). To identify symptoms that were mentioned in tweets, we applied two symptom extraction methods. Amazon Comprehend Medical tool was applied to an entire set of tweets. We considered two medical entities (symptoms and signs) as symptoms that users mentioned. We also implemented a symptom extraction model using Scispacy (ver 0.2.4). Scispacy is a Python package to handle scientific document and extracts medical and clinical terminology^7^. The model was trained on publicly available domain-specific corpus of medical notes which consists of 1,500 PubMed articles with over 10,000 disease and related chemical terms. The model identifies medical name entities in tweet texts. We considered the medical entity “disease” as a symptom that users mentioned. In total, 51 symptoms from 574 COVID-19 positive users were identified from both symptom identification methods.

### Association between clinical and demographic variables and clinical outcomes

To explore the consistency between individual variables and clinical outcomes, we defined three types of associations. Positive associations indicated that a variable in the outcome group showed a hazard/odd ratio >1, or higher value with a statistical significance (*p*-value < 0.05) compared to the control group. In case a statistical test was not performed, we decided that there was a positive association when the outcome group showed a 1.5-fold higher value than the control group. Negative association indicated that a variable in the outcome group showed a hazard/odd ratio <1, or lower value with a reported statistical significance (*p*-value < 0.05) compared to the control group. In case a statistical test was not performed, we decided that there was a negative association when the outcome group showed a 1.5-fold lower value than the control group. When there was no significant change between the outcome group and control group, no-association was assigned between variable and clinical outcome. In terms of sex, when there were more males compared to females, we assumed there was a positive association with an outcome based on the case studies of sex and age of COVID-19 patients in Italy^8^ and New York City^9^ (as of April 14, 2020). To identify outcome-specific clinical and demographic variables in biomedical literature, we performed consensus analysis. Seven outcomes which were studied at least twice, and 107 variables tested in ≥2 for a given outcome studies were used for further analyses. Variables showing positive associations in >50% of studies, which investigated same output, were defined as outcome-specific variables (risk factors).

## Results

### 1. Landscape of clinical and demographic variables of COVID-19 in biomedical literature

To understand the clinical and demographic properties of COVID-19, we systematically analyzed 45 recently published biomedical studies (**figure 1A** and **appendix 1**). The literature evaluated 299 clinical variables (92 comorbidities/complications, 124 laboratory findings, 49 treatment options, and 34 symptoms) and 4 demographic variables (age, sex, alcohol drinking history, and smoking history) in 13 clinical outcomes. Seven outcomes (disease severity, death, ICU admission, diagnosis, acute respiratory distress syndrome (ARDS), O_2_ saturation, and hospitalization) have been studied at least twice (**figure 1**); on average, each study examined 72 variables, and 102 variables were assessed in at least five studies. Age and sex were measured in more than 80% of the studies. Diabetes and hypertension were the most commonly measured comorbidities (>50% of studies). Fever, cough, myalgia/fatigue, chest tightness/dyspnea, diarrhea, and headache/dizziness were the most commonly measured symptoms (>50% of studies). Eleven laboratory test values that measured liver and kidney function (*e.g*., alanine aminotransferase and aspartate aminotransferase) and hematologic index (*e.g*., lymphocytes, platelets and neutrophils) were examined in >50% of studies. Therapy involving antiviral agents, antibiotics, and oxygen inhalation were used in more than 30% of the studies (**appendix 2**).

**Figure 1.**
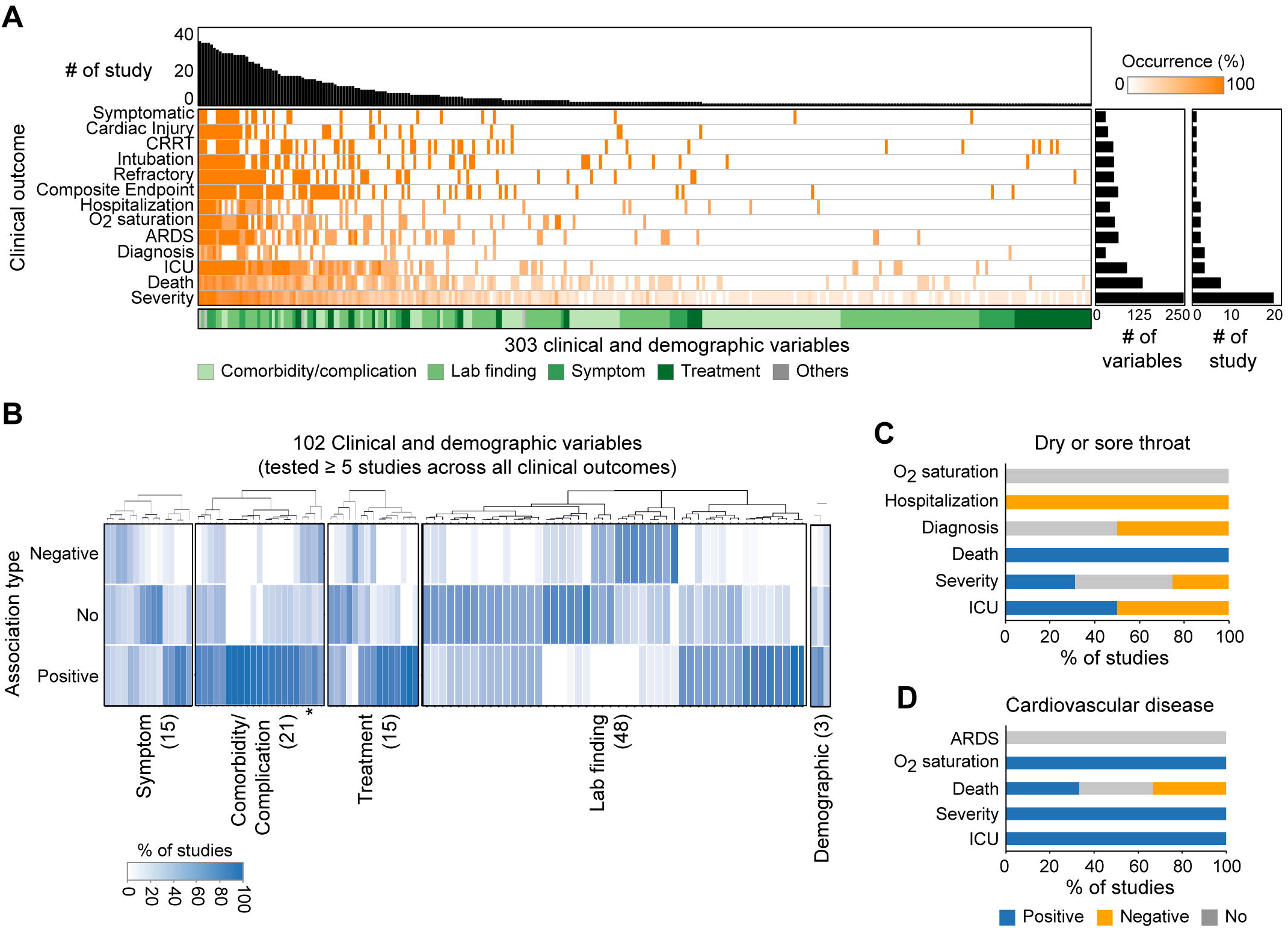
Properties of clinical and demographic variables of COVID-19. **(A)** Landscape of clinical and demographic variables. **(B)** Association between variables and overall clinical outcomes. Number of variables tested in ≥5 studies were placed in brackets. Asterisk indicated association types of cancer. (**C and D**) Association types of clinical outcomes of COVID-19. Association types of (**C**) dry or sore throat and (**D**) cardiovascular disease depending on different clinical outcomes were shown.

We next investigated the association between identified variables and clinical outcomes of COVID-19. We considered 102 frequently tested variables (≥ 5 studies), and examined the proportion of studies that showed positive-, negative-, and no-associations between clinical outcomes and a given variable (**appendix 3**). Positive associations indicated higher values (*e.g*., disease severity increases as patients get older), while negative associations indicated lower values (*e.g*., disease severity increases as basophil count gets lower) of variables associated with clinical outcomes (see **Methods** for details). No-association indicated there was no relation between variables and outcomes. We found that the majority of variables (95 variables, 93%) had inconsistent associations across clinical outcomes showing mixed association types (**figure 1B**). Of those, 46 (45%) variables had all three types of associations. For example, cancer showed positive-, negative-, and no-associations in 58%, 26%, and 16% of studies, respectively (**asterisk in figure 1B**). 43% (9/21) of comorbidity/complication, 40% (6/15) of treatment, and 73% (11/15) of symptom variables showed all three types of associations. Laboratory findings showed relatively more consistent associations with clinical outcomes: 38% (18/48) of variables showed all association types. Furthermore, we found that variables had unique association types depending on different clinical outcomes. Dry or sore throat, one known symptom of COVID-19, showed positive-, negative- and no-associations in death, hospitalization and O_2_ saturation, respectively (**figure 1C**). Meanwhile, it showed mixed associations with other clinical outcomes, such as disease severity, ICU admission, and diagnosis. Cardiovascular disease, one common comorbidity of COVID-19, showed mixed associations in death, positive association with ICU and disease severity, but no association with ARDS (**figure 1D**).

### 2. Consensus identification of outcome-specific clinical and demographic variables

Our previous observations suggested that at the time of publication, there were no effective treatment options, or well-identified symptoms, comorbidities, and lab findings to predict outcomes of COVID-19. Therefore, it seemed relevant to find a set of clinical and demographic variables that were specific for individual outcomes for better guidance of disease detection, treatment, and control. To generalize the importance of clinical and demographic variables, a consensus (level of agreement) analysis was performed. We collected variables that were tested at least twice in a given outcome and defined them as outcome-specific variables when they showed positive associations with a given outcome in more than half of studies (**figure 2** and **appendix 4**). We observed that different sets of variables were associated with individual outcomes, meanwhile, no variable was universal across all clinical outcomes (**figure 2A**). Age was specific variable for ARDS, disease severity, death, ICU admission, and O_2_ saturation, but not specific for diagnosis and hospitalization (**figure 2B**). Sex (male) was specific variable for ARDS, disease severity, death and ICU admission. Three lab findings (D-dimer, C reactive protein and lactate dehydrogenase) were specifically associated with four outcomes. Diabetes and hypertension (comorbidity/complication) were specific for disease severity and death. Also, there were variables only specific for one outcome. Arrhythmia, thyroid disease (comorbidity/complication), confusion/fluster, tonsil swelling, enlargement of lymph nodes/sinus (symptom), and levels of IL-10 and NT-proBNP (N-terminal-pro hormone BNP, lab finding) were only specific for the severity of disease progression. Level of prothrombin time was specific variable for ICU admission. For death, SOFA score (lab finding), and anemia (symptom) showed positive associations. Fever was a specific variable for O_2_ saturation. There was no significantly associated variable for diagnosis and hospitalization indicating the current lack of clinical understanding at the early stage of COVID-19. Identified outcome-specific variables could be surrogate risk factors to identify COVID-19 patients and determine their treatment options.

**Figure 2.**
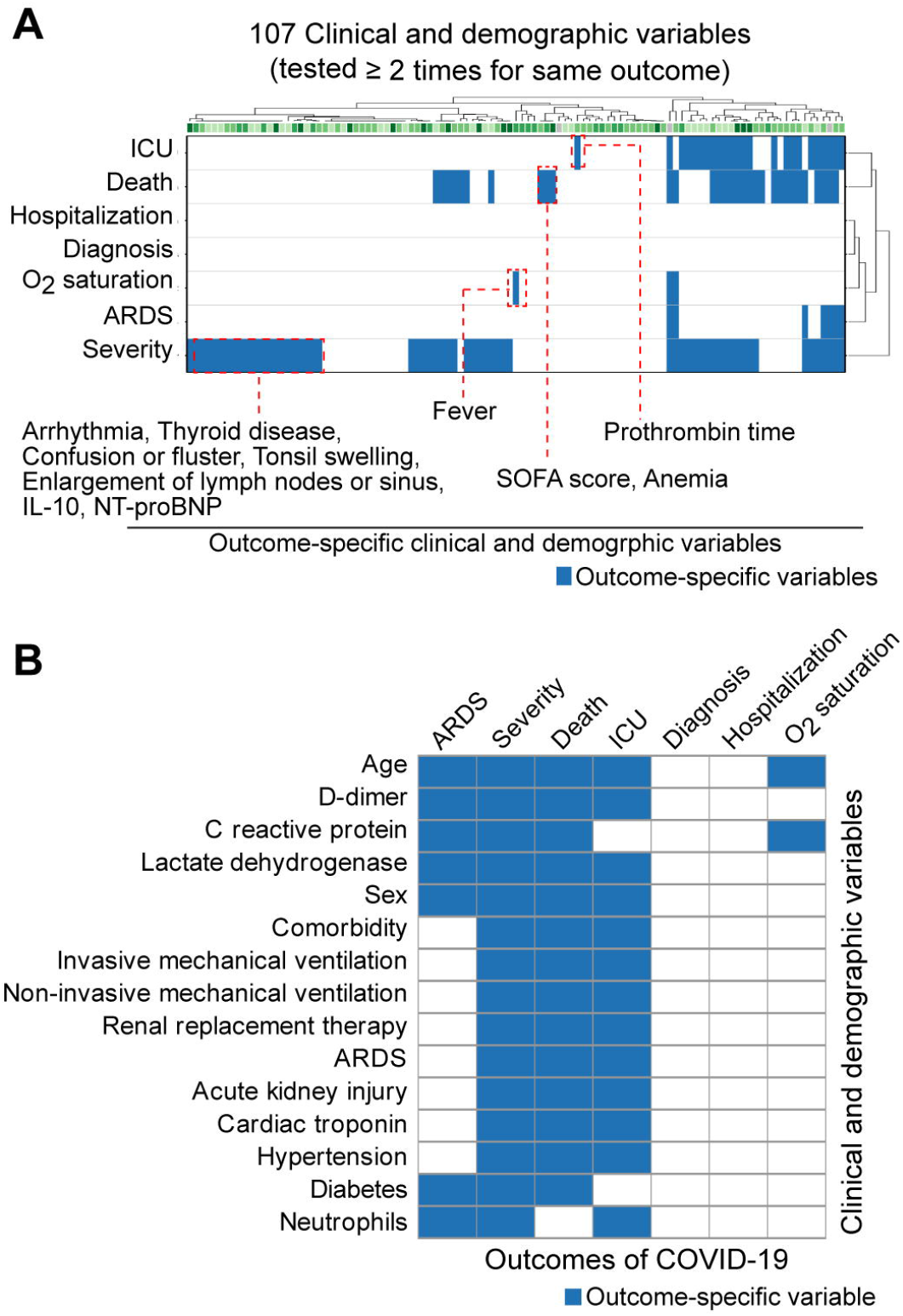
Consensus identification of outcome-specific clinical and demographic variables. (**A**) Outcome-specific clinical and demographic variables in a given outcome of COVID-19. Variables that were only specific for one clinical outcome were shown in red dashed box. Clinical and demographic variables that were specific for at least three outcomes were presented at (**B**). Blue indicated identified outcome specific variables.

### 3. Expanding the COVID-19 symptom landscape and identifying novel symptoms using social media

Early identification of symptoms is important for successful treatment of disease^10^. Although COVID-19 showed heterogeneous and uncharacterized symptoms, biomedical literature considered a limited number of symptoms known for infectious diseases, such as fever, cough, and fatigue. Social media can provide rapid and efficient surveillance of disease risk and outbreaks^11,12^. Therefore, we decided to expand the symptom landscape by integrating social media data with biomedical literature. We first identified 1,036 twitter users who introduced themselves as COVID-19 positive patients, and selected 574 users (55%) who openly and voluntarily discussed their COVID-19 symptoms (see **Methods** for details). In total, 51 symptoms were identified in social media data (**figure 3A** and **appendix 5**). On average, individual users mentioned 3 different symptoms (range of 1-15). We grouped 51 symptoms into three categories based on their frequency of mention (**figure 3B** and **appendix 5**). Common symptoms (11 symptoms) were mentioned by >10% of users. Many were non-specific symptoms of respiratory infections, such as fever, cough, and chest tightness/dyspnea. However, 14 symptoms were potentially COVID-19-specific, and rarely reported in social media (rare, <1% of users). Sputum, dehydration, enlargement of lymph nodes or sinus, and oral problems (*e.g*., abrasions in mouth, mouth ulcers, sensitive teeth, toothache, and dry mouth) were defined as rare symptoms. Twenty-six symptoms were observed ranging from 1% to 10% of users (less common). They included chills, chest pain, gastrointestinal symptoms, and skin problems such as blister, dry skin, chapped lips, and itching. We next examined the symptoms that were mentioned together (**figure 3C** and **appendix 6**). Of 612 symptom pairs that co-occurred, 264 (43%) were between common and less common symptoms (**figure 3D**). One major cluster of symptoms was frequently mentioned together (**figure 3E**). They were composed of 8 common (dry or sore throat, fever, chest tightness/dyspnea, cough, weakness, myalgia/fatigue, headache/dizziness, and body ache/pain: neck and back pain, and general body ache) and 8 less common symptoms (cold-like symptoms, chest pain and congestion, gastrointestinal symptoms, chills, stuffy or runny nose, nausea/vomiting, and respiratory symptoms, such as lung pain). There were also two pairs of rare symptoms (**figure 3D**): anemia-weight gain and anemia-urination problems (*e.g*., urinary retention and weakened bladder).

**Figure 3.**
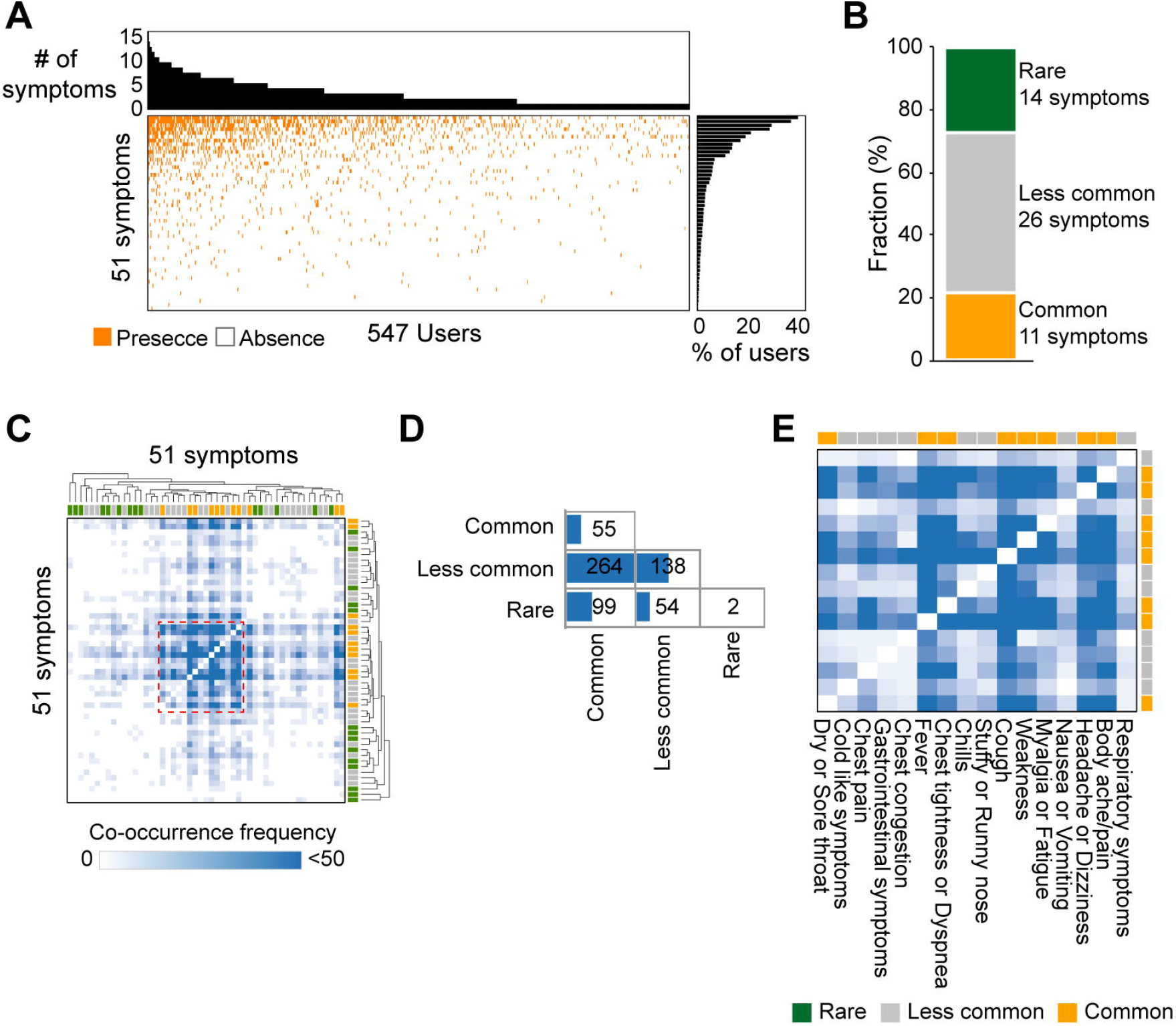
Covid-19 related symptoms extracted from social media data. **(A)** Landscape of symptoms identified from social media data. Orange and white indicate the presence and absence of symptoms in a given user, respectively. **(B)** Fraction of common, less common and rare symptoms. Common symptoms were mentioned from >10% of users, and rare symptoms were mentioned from <1% of users. **(C)** Co-occurrence of symptoms. One major cluster was shown in red dashed box. **(D)** Number of symptoms pairs depending on mentioning frequency. Blue bars (bottom) indicate the number of co-occurred pairs. **(E)** One major cluster of symptom pairs. Green, grey, and orange indicate rare, less common and common symptoms, respectively.

Finally, we identified novel symptoms potentially related to COVID-19. From the integration of literature and social media, we identified 59 symptoms (**figure 4** and **appendix 7**). Among them, 25 (42%) were novel symptoms that were only mentioned in social media, but were not considered in the literature. Loss of smell or taste, and body ache/pain were frequently mentioned common symptoms in social media. Eye-(*e.g*. dry eye and eye pain) and ear-(*e.g*. ear pain and ear ache) problems, sweating, sneezing, and allergy-like symptoms were mentioned at a moderate frequency only in social media. Of 14 rare symptoms, 10 were only observed in social media. They included oral problem, hair loss, urination problem, and dehydration. These social-media-specific rare symptoms would be potential novel symptoms for COVID-19, when we considered that 4 rare symptoms (abdominal pain, anemia, sputum, and enlargement of lymph nodes or sinus) were already evaluated in the literature (**figure 4**).

**Figure 4.**
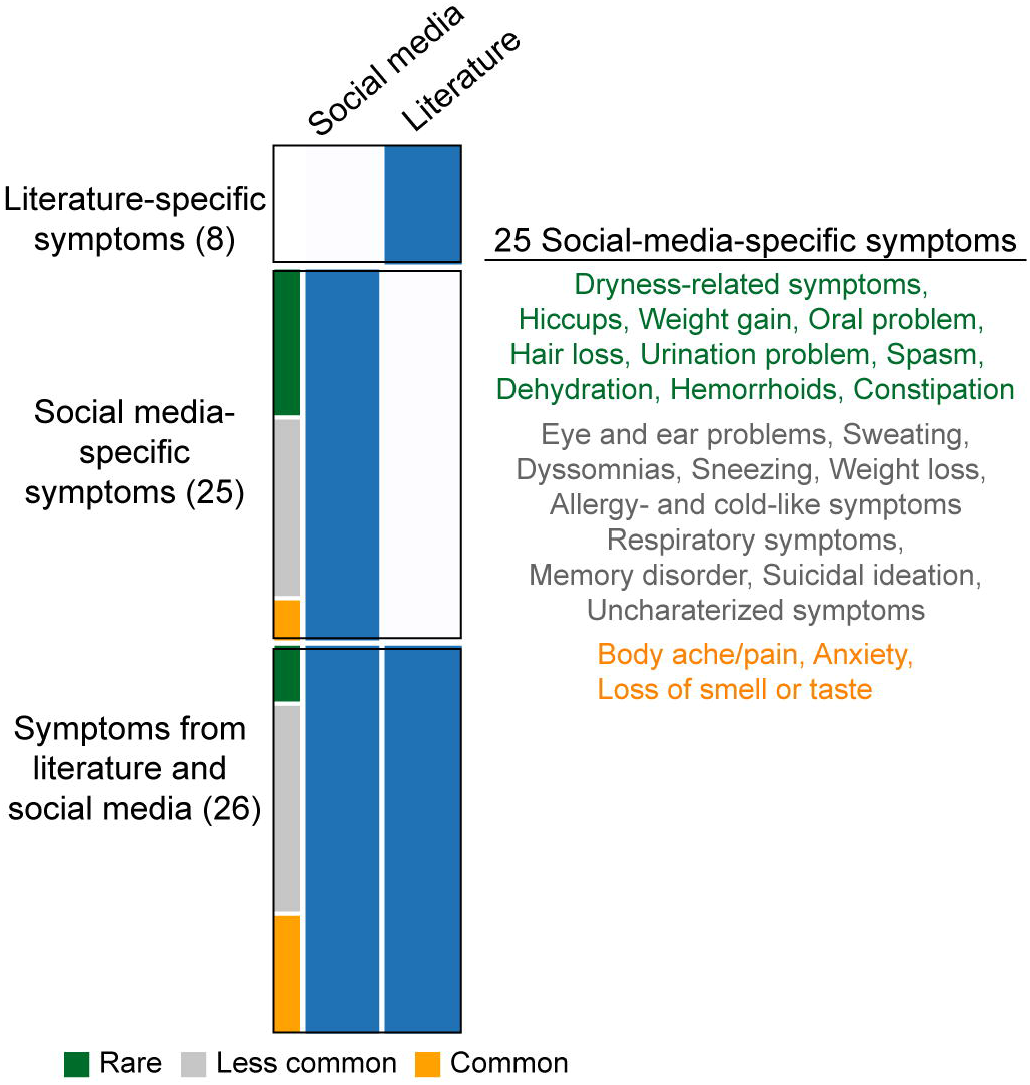
Identification of novel symptoms of COVID-19. Comparison of symptoms between biomedical literature and social media data. Symptoms that were observed in literature or social media were colored in blue. 25 social-media-specific symptoms were presented. Green, grey, and orange indicated rare, less common, and common symptoms, respectively.

## Discussion

Our study showed inconsistency of clinical and demographic variables across clinical outcomes. From the consensus analysis, we identified outcome-specific clinical and demographic variables that may be helpful to find patients and predict their specific clinical outcomes providing appropriate treatments. From the integration of biomedical literature and social media data, we expanded the repertoire of symptoms. In addition to more commonly reported symptoms of COVID-19 (*e.g*., fever, fatigue, and chest tightness), we identified loss of smell or taste, body ache and back/neck pain, as well as other less common and rare symptoms. Indeed, loss of smell or taste was recently proposed as one of the key features of a COVID-19 diagnosis model^13^.

In social media data, we observed that certain combinations of symptoms were frequently observed among COVID-19 patients. Interestingly, we identified two pairs composed of rare symptoms (anemia -weight gain and -urination problems, such as urination retention and weakened bladder). It has been shown that COVID-19 attacks hemoglobin in red blood cells and restricts oxygen transportation^14^. A persistent reduction of oxygen transportation leads to the development of anemia^15^. Urinary bladder is enriched with ACE2 positive cells and proposed as a target organ for COVID-19 invasion^16^. Our results highlighted that combinations of symptoms would guide the reliable identification of COVID-19 patients rather than a single common symptom which possibly identifies false positives.

One of the limitations of our study is the self-reported nature of social media data and the lack of more detailed information from the patients. We observed that 55% of social media users who were positive for COVID-19 mentioned symptoms, and 8% mentioned potential comorbidities. Thus, only 63% of social media users indicated any form of COVID-19 conditions, which means that at least 37% of users could be either false positives (they were not COVID-19 positive users) or asymptomatic patients. Alternatively, it is possible that we have not captured all of the COVID-19 positive patients in our social media collection due to the limited amount of keyword searches. Nevertheless, various articles have indicated that between 4% and 78% of all COVID-19 positive patients were asymptomatic^17^, and this seems to vary widely based on age of patients, test location, and time of testing after infection^18–21^. Thus, our research is in line with other studies demonstrating the vast range of COVID-19 patients who show or report no symptoms. It should also be noted that Twitter was the source of social media data that we examined, and perhaps more symptoms would be discovered if we analyzed other various sources. Twitter does have a wide, representative user base around the world, and provides open source information that can be easily gathered, but future research could examine alternate social media sources.

Although social media may lack depth of patient information, it provides an effective method of collecting breadth of data. Social media data can be gathered non-invasively across the world, 24 hours a day, and is an extremely efficient method^22^ for rapidly disseminating new knowledge related to COVID-19. In other words, clinicians and scientists can collect new patient information from various regional locations, as well as quickly circulate public service announcements for a wide range of audiences. Social media hubs provide a useful alternate source of patient data to explore clinical characteristics of various disease states and populations.

Another limitation of our research involves the limited number of available biomedical literatures of individual outcomes. Of 13 reported clinical outcomes for COVID-19, 7 were studied at least two times, limiting opportunities to perform more systematic and consensus analyses of the landscape of risk factors and symptoms. Use of additional literature that will be published in future and electronic health record (EHR) studies^23^ may refine the assessment of risk factors and symptoms, and increase the accuracy of patient identification for different clinical outcomes.

In conclusion, our current research demonstrates the extensive variability of COVID-19 risk factors and symptoms, and the usefulness of utilizing alternative sources of patient data (*i.e*., social media) to uncover less common or other types of symptoms that are not reported in clinical studies. Our findings show the practicality and feasibility of employing social media data for investigating new disease states. These practices could be incorporated into routine procedures for early COVID-19 identification, and the determination of their clinical outcomes, providing appropriate interventions and treatments.

## Data Availability

All identified clinical and demographic variables from biomedical literature and their association types with outcomes of COVID-19 are available in Appendix 2 and 3. Potential risk factors and critical symptoms that are specific for different outcomes are available in Appendix 4. A list of symptoms that were mentioned in social media and their frequency (fraction of users) are available in Appendix 5. Co-occurrence of symptoms are tabulated at Appendix 6. A list of novel symptoms of COVID-19 is available in Appendix 7.

## Contributors

J.J., and G.B. designed the study. J.J., G.B., and S.S. collected the Data. J.J., and A.P. analyzed the data. J.J. interpreted the data, and wrote the manuscript with contributions from A.P. All authors revised the manuscript and contributed to the final review and editing, and have approved the final manuscript.

## Data sharing

All identified clinical and demographic variables from biomedical literature and their association types with outcomes of COVID-19 are available in Appendix 2 and 3. Potential risk factors and critical symptoms that are specific for different outcomes are available in Appendix 4. A list of symptoms that were mentioned in social media and their frequency (fraction of users) are available in Appendix 5. Co-occurrence of symptoms are tabulated at Appendix 6.

## Acknowledgements

We would like to thank all members of the Labs team at Klick Inc. for their helpful advice and assistance. We especially thank Maheedhar Kolla, Jeremy J. Jurksztowicz, JJ Mifsud, and Michael Lieberman for technical support and scientific advice for the study.

